# HIV escape and resistance in the central nervous system in treatment experienced South Africans

**DOI:** 10.1101/2023.03.03.23286708

**Authors:** Dami Collier, Anne Derache, Farina Karim, Theresa Smit, John Adamson, Khadija Khan, Tasneem Naidoo, Nirmala Perumal, Jay Brijkumar, Jennifer Giandhari, Tulio De Oliveira, Alex Sigal, Steve Kemp, Ravindra K. Gupta, HERB Study Team

## Abstract

HIV associated neurocognitive disorder (HAND) remains an important HIV-associated comorbidity despite antiretrovirals (ARVs). Cerebrospinal fluid (CSF) escape/discordance is now recognised in the context of individuals with a reconstituted immune system with an estimated prevalence of 10%. However, the contribution of CSF escape/discordance to HAND remains uncertain. Furthermore, a latent reservoir of HIV in the brain has implications for lasting cure strategies. Little is known about the prevalence of CSF escape/discordance amongst people living with HIV (PLWH) in sub-Saharan Africa (SSA).

We conducted a longitudinal cohort study of PLWH who were 18 years or older on ARVs for at least 1 year who reported neurocognitive complaints. We obtained paired CSF and blood at baseline, 6, 12 and 24 months. Viral load (VL) testing was done with the Abbott m2000 RealTime System. HIV genotyping was done by Sanger sequencing and next generation sequencing (NGS) by Illumina MiSeq. Resistance calling was done using Stanford HIV drug resistance database. Random drug levels were done on plasma and CSF using mass spectrometry.

We present the results at baseline. Seven hundred and eight adult PLHIV attending a HIV treatment centre were screened using the Simioni symptom questionnaire and in addition asked “do you have a chronic, persistent headache?” Fifty-nine PLHIV answered yes to at least one of the screening questions and were considered for enrolment. Thirty consented to participate. The median age was 37.6 (IQR 33.2 to 48.3) years. The majority were women (98.0%, 28/30). Headache was the most common symptom (93.3%), then memory impairment (56.4%), attention deficit (48.3%) and impairment in executive functioning (46.7%). All participants had Karnofsky performance scale > 70% and were able to perform their activities of daily living independently. Symptoms of depression were common, with 82.8% scoring a CESD-R-10 >10. The median duration of ART was 9.9 (IQR 5.7 to 11.9) years. 72.4% (n=21/29) were on tenofovir/emtricitabine/efavirenz. The rest were on second line ARVs (ritonavir boosted lopinavir plus zidovudine/lamivudine or tenofovir/emtricitabine). The median nadir CD4 count was 193 (IQR 98 to 301) cells/mm^3^ and the current median CD4 count was 547 (IQR 384 to 856) cells/mm^3^. At baseline 86.2% had an undetectable plasma HIV viral load (<40 copies/ml) (25/29). Eighteen participants had paired CSF and blood successfully sampled. Of these 4 had detectable virus in the blood with VL ranging from 82 to 38,992 copies/ml. Two participants had CSF escape/discordance (9.0% (2/22) and 2 others had detectable VL in CSF but lower than the VL in blood. We found an association between detectable CSF VL and viraemia (p 0.001) and boosted protease inhibitor (PI) based ART (p 0.02). All participants who had undetectable VL in blood and CSF had detectable blood and CSF drugs levels corresponding to their prescribed ARVS. Two participants with detectable VL in the CSF had drug levels measured, which was below the limit of detection of the drug assays in both blood and CSF. Both participants were on second line boosted PI based ARVs. Viral sequencing revealed NNRTI resistance mutation G190A detected in both blood and CSF in the participant with CSF discordance and NRTI M184V, NNRTI K103N and P225H in the blood but not CSF of the second participant. No minority variants were identified below 20% by NGS.

Virological failure appears to be driven by poor treatment adherence. The results suggest that the prevalence of CSF escape/discordance in HIV positive neurosymptomatic persons is consistent with previously published prevalence from resource rich settings however this needs to be explored in the larger study. Symptoms of depression were common and may bias self-reported neurocognitive impairment and needs to be explored further. This ongoing longitudinal study will also investigate the evolution of drug resistant variants in CSF and the relationship with plasma viral quasispecies.

## Introduction

The brain is considered an anatomically privileged site and HIV-1 entry to the central nervous system (CNS) is facilitated by loss of the integrity of the blood brain barrier (BBB) early in HIV infection. HIV is believed to ingress into the CNS as free particles or within activated CD4 central memory T cell or monocytes and establish a latent reservoir. The viral reservoir is thought to be maintained by clonal expansion of latently infected T cells or low-grade independent replication in myeloid lineage cells such as macrophages or microglia(1-3) and poor CNS drug penetration could allow HIV-1 replication to continue. This is a barrier to achieving lasting eradication of HIV from active and latent reservoirs and is a research priority for HIV cure efforts.

CNS HIV-1 replication has been associated with pleocytosis and abnormal MRI appearances(4), suggestive of neuronal damage and neuroinflammation and has been implicated in reported neurocognitive impairment in people living with HIV (PLWH) despite suppressive combination antiretroviral therapy (ART)(5-7). So-called cerebrospinal fluid (CSF) escape/discordance is now recognised in the context of individuals with a reconstituted immune system(8, 9) and may occur in up to 10% of individuals(10). Recent studies have shown evidence of compartmentalised HIV-1 CSF viruses with resistance associated mutations (RAMs)(11-14) and in those with unexplained low level viraemia (LLV)(15). This may be explained by ongoing replication of HIV in CNS with resultant seeding to plasma.

The contribution of HIV CSF escape/discordance and to neurocognitive impairment remains poorly understood in sub-Saharan Africa (SSA)(16). This study aimed to determine the prevalence of CSF viral escape/discordance in South African patients with mild-moderate chronic neurological symptoms, to determine if these viruses are compartmentalised and to track the evolution of RAMs in CNS compartmentalised HIV-1 in ART experienced PLWH.

## Method

### Design

This paper summarises the pilot phase of the ongoing longitudinal cohort study of CSF escape and the evolution of drug resistance in a cohort of neurosymptomatic HIV positive persons on long term ART in Durban, South Africa.

### Setting

Durban eThekwini district, KwaZulu-Natal, South Africa.

### Participants

PLWH who are 18 years and older on ART treatment for at least 1 year who present at any of the Durban hospitals and require a lumbar puncture (LP) for clinical reasons, including but not limited to neurocognitive complaints (memory difficulties, poor concentration, ineffective learning and difficulties in decision making or executive function); headache; fever; evidence of DWMSA on brain MRI.

### Exclusion criteria

Those with coma, seizure or overt CNS diseases of infectious aetiology (such as TB, cryptococcal or bacterial meningitis) or CNS malignancy determined through a radiological or laboratory diagnosis; a combination of CSF polycythaemia with WCC greater than 5 cells/μL, CSF glucose less than 60% of serum glucose and protein greater than 50 mg/dL; and peripheral blood contamination of CSF with RBC greater than 1000 cells/μL.

### Protocol summary

Eligible participants who gave their informed consent were recruited and underwent a baseline assessment including a detailed questionnaire collecting demographic and clinical data including; CD4, viral load (VL), clinical course, ART history, an International HIV dementia screen (IHDS), Karnofsky performance scale to measure functional impairment, Center for Epidemiologic Studies Depression Scale Revised (CESD-R-10) to screen for depression and the Alcohol Use Disorders Identification Test (AUDIT) to assess alcohol consumption. ART adherence was determined by the ACTG Adherence tool. Paired CSF and blood specimens were collected. Plasma aliquots were prepared by centrifuging whole blood specimens at 800 x g for 10mins and aliquots stored at -80°C. CSF was centrifuged at 800 x g for 10mins and both supernatant aliquots and the cell pellets were stored. Peripheral blood mononuclear cells were harvested and stored in aliquots at - 80°C. Random or trough ART drug levels were measured using tandem liquid chromatography-mass spectrometry on paired CSF and blood. Brain MRI was performed at 3 Tesla field strength; protocol included volumetric T1- and T2-weighted sequences, fluid-attenuated inversion recovery (FLAIR), and diffusion tensor imaging with at least 6 diffusion gradient directions with intravenous gadolinium contrast given when clinically necessary. Images were reviewed in real time for clinical safety and quality control by a local radiologist. Follow up was planned at 6, 12 and 24 months with repeat paired CSF and blood sampling should neurocognitive symptoms persist.

### Viral load testing

Performed on paired CSF and blood using the Abbott m2000 RealTime System™ with HIV-1 VL determination from human plasma of HIV-1 infected individuals in the range of 40 to 10 000 000 copies/ mL (Abbott Molecular Inc., Des Plaines, IL, USA).

HIV genotyping and resistance testing-Detectable HIV in blood and CSF was amplified and sequenced by Sanger population sequencing and whole genome sequencing was done using the Illumina Miseq platform. Resistance calling was done using Stanford HIV drug resistance database(17). Multiple sequence alignment using consensus sequences was performed using using mafft v7.515(18). Phylogenies were inferred with IQTREE2 v2.2.2(19) using a GTR+F+R6 model with 1000 rapid bootstraps.

### Ethics

Approval was granted by UKZN Biomedical Research Ethics Committee BE604/17 and the University College Hospital Research Ethics Committee 14865/001.

## Results

Between April 2018 and March 2020, 708 adult PLWH attending a HIV treatment centre were screened using the Simioni symptom questionnaire and in addition asked “do you have a chronic, persistent headache?” No participant had a prior brain MRI. 8.3% (59/708) of screened patients reported yes to any of the 4 questions. Thirty of these were consented to participate and were recruited (Figure 1). The study was paused due to the global COVID-19 Pandemic in March 2020.

**Figure 1:**
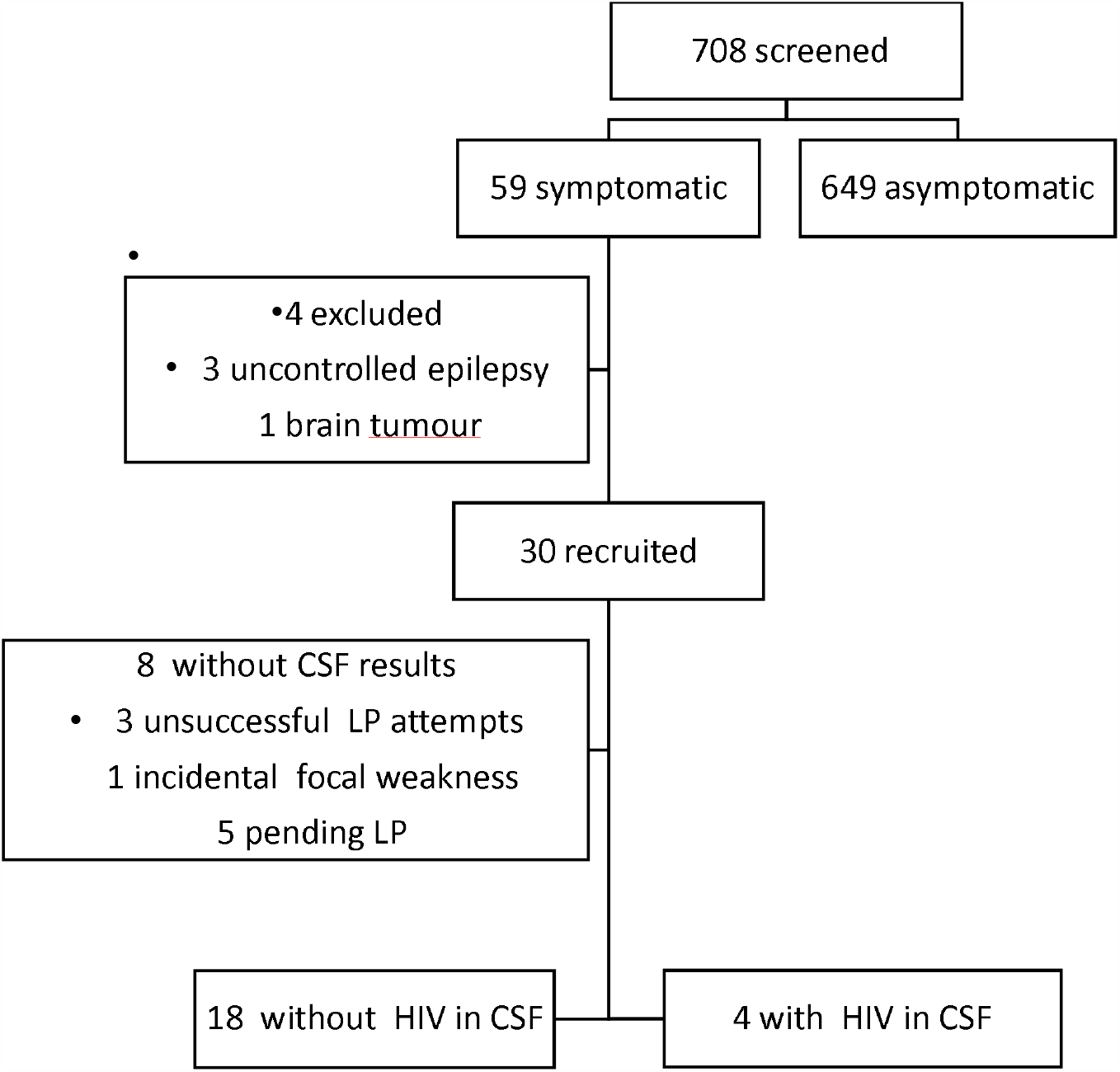
study flow diagram

Table 1 summarises characteristics of the cohort. The median age was 37.6 (IQR 33.2 to 48.3) years. The majority were women (93.3%, 28/30). Headache was the most common symptom (93.3%), followed by memory impairment (56.7%), impairment in executive functioning (46.7%) and attention deficit (48.3%). All participants had Karnofsky performance scale > 70% and were able to perform their activities of daily living independently. Symptoms of depression were common, with 82.8% scoring a CESD-R-10 > 10. The median duration of ART was 9.9 (IQR 5.7 to 11.9) years. 72.4% (n=21/29) were receiving ART combination therapy (tenofovir+emtricitabine+efavirenz/nevirapine). The rest were receiving second line ART (ritonavir boosted lopinavir plus zidovudine/lamivudine or tenofovir/emtricitabine). The median nadir CD4 count was 193 (IQR 98 to 301) and the current median CD4 count was 547 (IQR 384 to 856). At baseline 86.2% (25/29) had an undetectable plasma HIV viral load of <40 copies/ml. Four participants had detectable virus in the blood with VL ranging from 82 to 38,992 copies/ml.

**Table 1:**
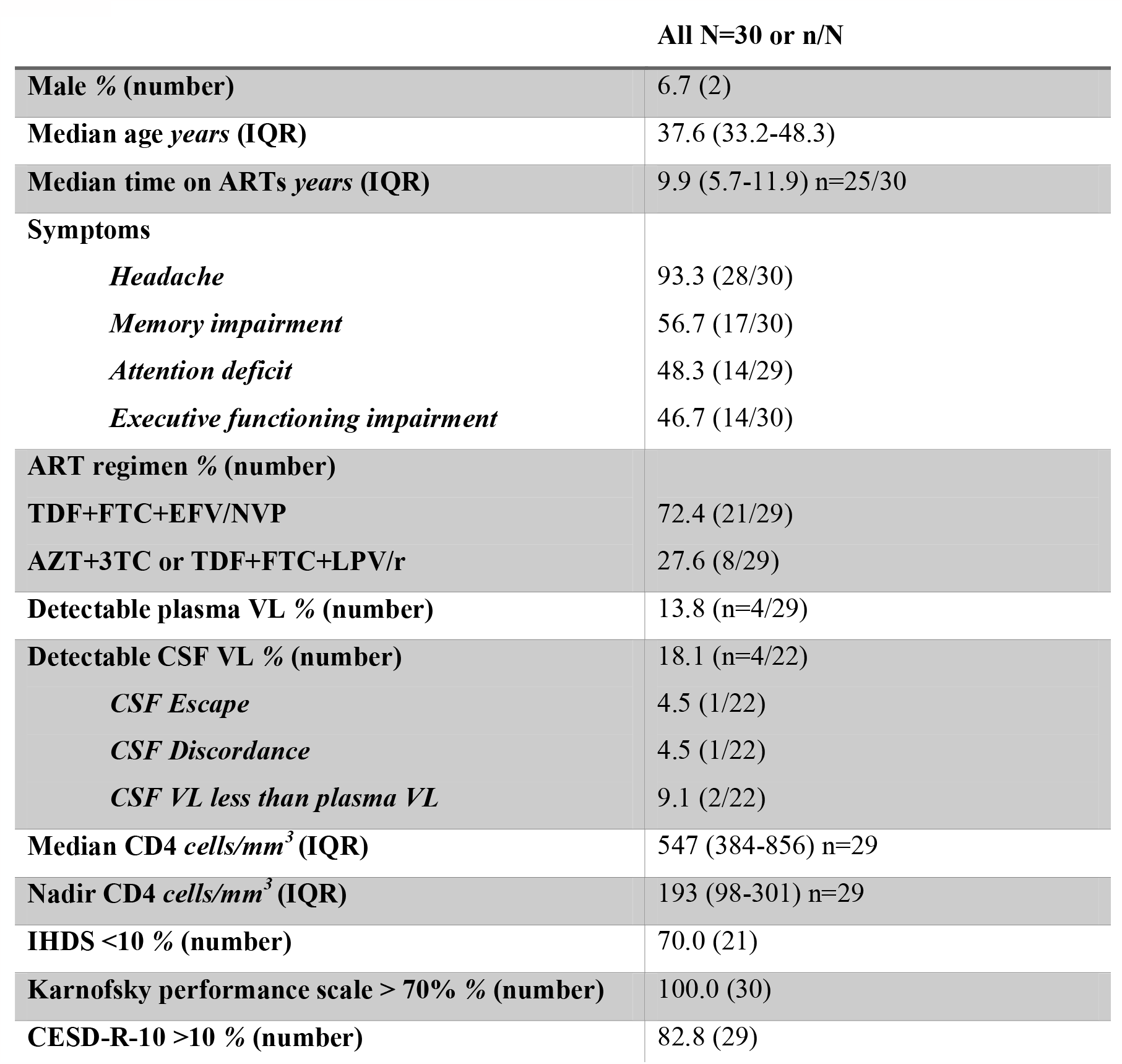
Study population description. IQR; interquartile range, ART; antiretroviral treatment, TDF; tenofovir, FTC; emtricitabine, EFV; efavirenz, NVP; nevirapine, AZT; zidovudine, 3TC; lamivudine, LPV/r; ritonavir boosted lopinavir, VL; viral load, CSF; cerebrospinal fluid, IHDS; international HIV dementia score, CESD-R-10; centre for epidemiologic studies depression scale-revised.

### CSF replication

Twenty-two participants had successful lumbar punctures (LP) performed alongside blood sampling. Four participants had detectable HIV-1 VL in their CSF, ranging from 57 to 30,180 copies/ml (Table 3). Three of these individuals also had detectable HIV in their blood (Table 3). One participant had CSF escape with undetectable VL in the blood but detectable VL of 57 copies/ml in their CSF. One participant had CSF discordance with CSF VL > 1 log_10_ of the blood VL and 2 others had detectable VL in the CSF but less than that in blood. The remaining 18 participants with LP performed showed HIV-1 VL below limit of detection in both blood and CSF. The association between detectable CSF HIV-1 VL and clinical and demographic factors were explored in a univariable analysis (Table 2). There was an association between detectable CSF HIV-1 VL with having a detectable plasma HIV-1 VL (p 0.001) and current treatment with boosted protease inhibitor (PI) based second line ART (p 0.02).

**Table 2:**
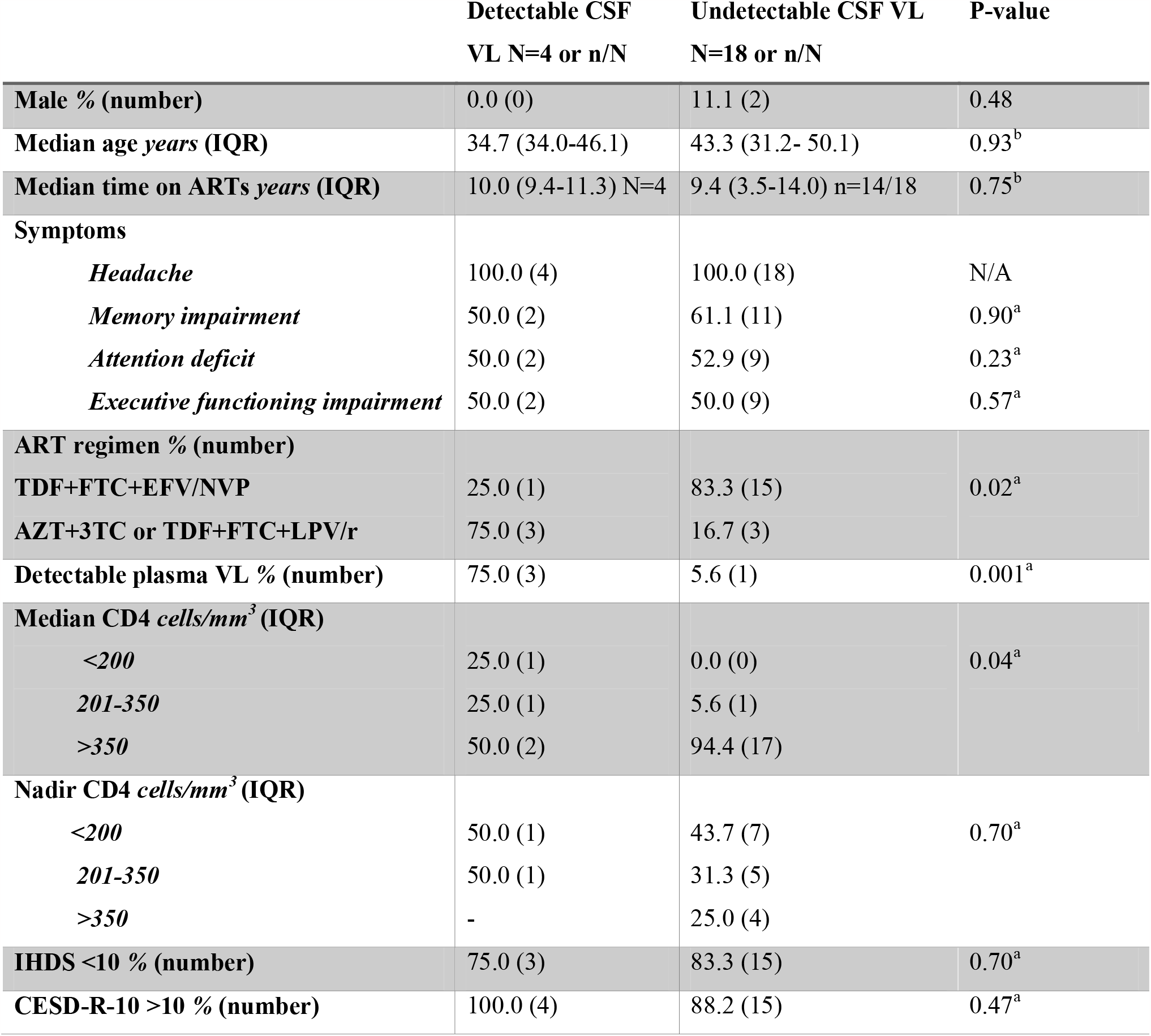
Association between detectable CSF viral load and clinical and demographic variables. ^a^ Chi-square test, ^a^ Mann-Whitney test, IQR; interquartile range, ART; antiretroviral treatment, TDF; tenofovir, FTC; emtricitabine, EFV; efavirenz, NVP; nevirapine, AZT; zidovudine, 3TC; IHDS; international HIV lamivudine, LPV/r; ritonavir boosted lopinavir, VL; viral load, CSF; cerebrospinal fluid, N/A; not applicable, dementia score, CESD-R-10; centre for epidemiologic studies depression scale-revised.

|  | Detectable CSF<br>VL N=4 or n/N | Undetectable CSF VL<br>N=18 or n/N | P-value |
| --- | --- | --- | --- |
| **Male % (number)** | 0.0 (0) | 11.1 (2) | 0.48 |
| **Median age years (IQR)** | 34.7 (34.0-46.1) | 43.3 (31.2- 50.1) | 0.93<sup>b</sup> |
| **Median time on ARTs years (IQR)** | 10.0 (9.4-11.3) N=4 | 9.4 (3.5-14.0) n=14/18 | 0.75<sup>b</sup> |
| **Symptoms** |  |  |  |
| *Headache* | 100.0 (4) | 100.0 (18) | N/A |
| *Memory impairment* | 50.0 (2) | 61.1 (11) | 0.90<sup>a</sup> |
| *Attention deficit* | 50.0 (2) | 52.9 (9) | 0.23<sup>a</sup> |
| *Executive functioning impairment* | 50.0 (2) | 50.0 (9) | 0.57<sup>a</sup> |
| **ART regimen % (number)** |  |  |  |
| **TDF+FTC+EFV/NVP** | 25.0 (1) | 83.3 (15) | 0.02<sup>a</sup> |
| **AZT+3TC or TDF+FTC+LPV/r** | 75.0 (3) | 16.7 (3) |  |
| **Detectable plasma VL % (number)** | 75.0 (3) | 5.6 (1) | 0.001<sup>a</sup> |
| **Median CD4 cells/mm<sup>3</sup> (IQR)** |  |  |  |
| <200 | 25.0 (1) | 0.0 (0) | 0.04<sup>a</sup> |
| 201-350 | 25.0 (1) | 5.6 (1) |  |
| >350 | 50.0 (2) | 94.4 (17) |  |
| **Nadir CD4 cells/mm<sup>3</sup> (IQR)** |  |  |  |
| <200 | 50.0 (1) | 43.7 (7) | 0.70<sup>a</sup> |
| 201-350 | 50.0 (1) | 31.3 (5) |  |
| >350 | - | 25.0 (4) |  |
| **IHDS <10 % (number)** | 75.0 (3) | 83.3 (15) | 0.70<sup>a</sup> |
| **CESD-R-10 >10 % (number)** | 100.0 (4) | 88.2 (15) | 0.47<sup>a</sup> |

**Table 3:**
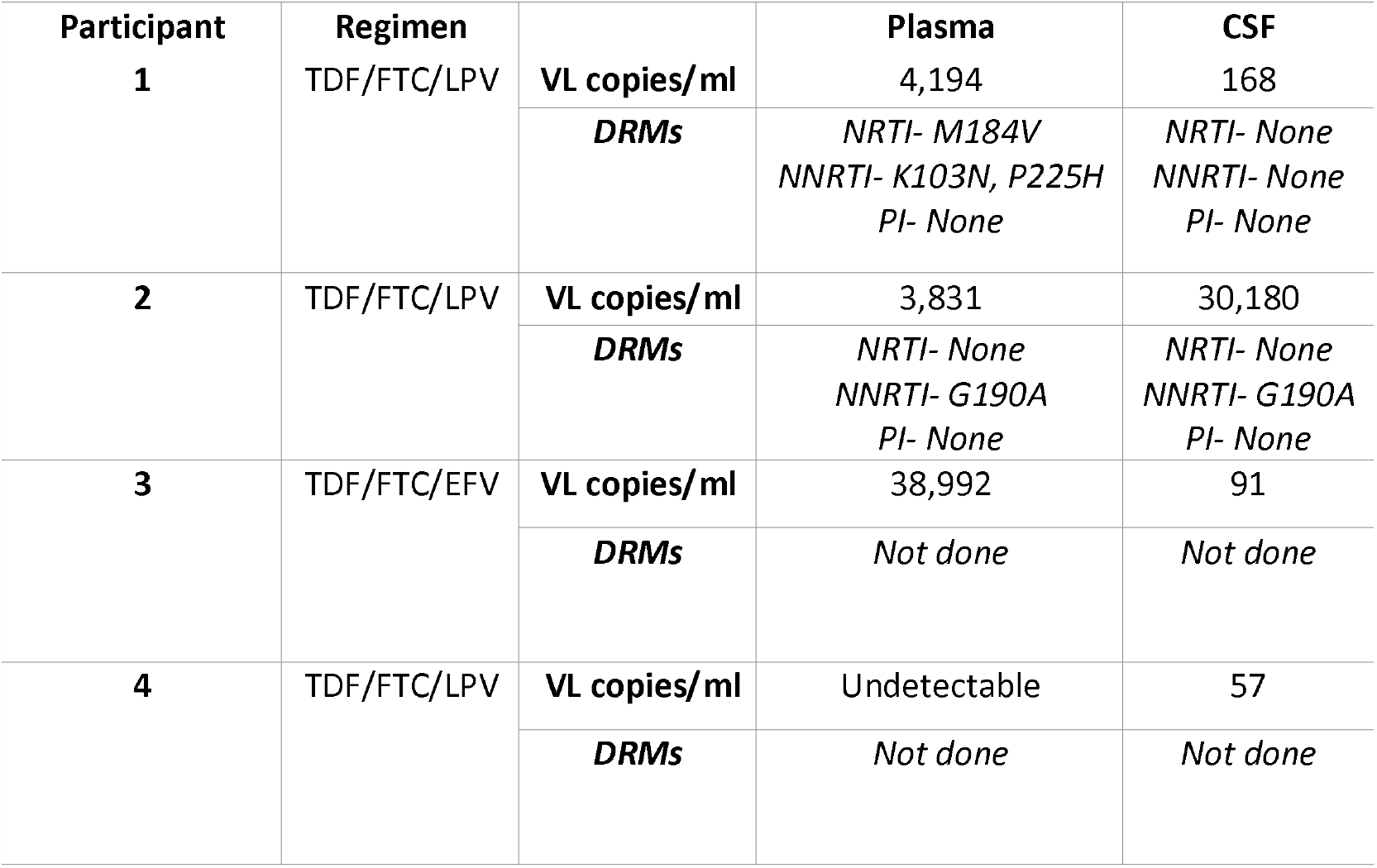
Fours participants with detectable HIV-1 in CSF at baseline. Showing viral load (VL) and drug resistant mutations (DRMs) if sequenced. TDF; tenofovir, FTC; emtricitabine, EFV; efavirenz, NVP; nevirapine, AZT; zidovudine, 3TC; lamivudine, LPV/r; ritonavir boosted lopinavir.

### Drug resistance

Viral sequencing revealed the NNRTI resistance mutation G190A detected in both blood and CSF in participant 2 with CSF discordance. Mutations conferring resistance to NRTI (M184V), and NNRTI (K103N and P225H) were observed in the blood but not CSF of participant 1 (Table 3 and Figure 2). No minority variants were identified below 20% by NGS.

**Figure 2:**
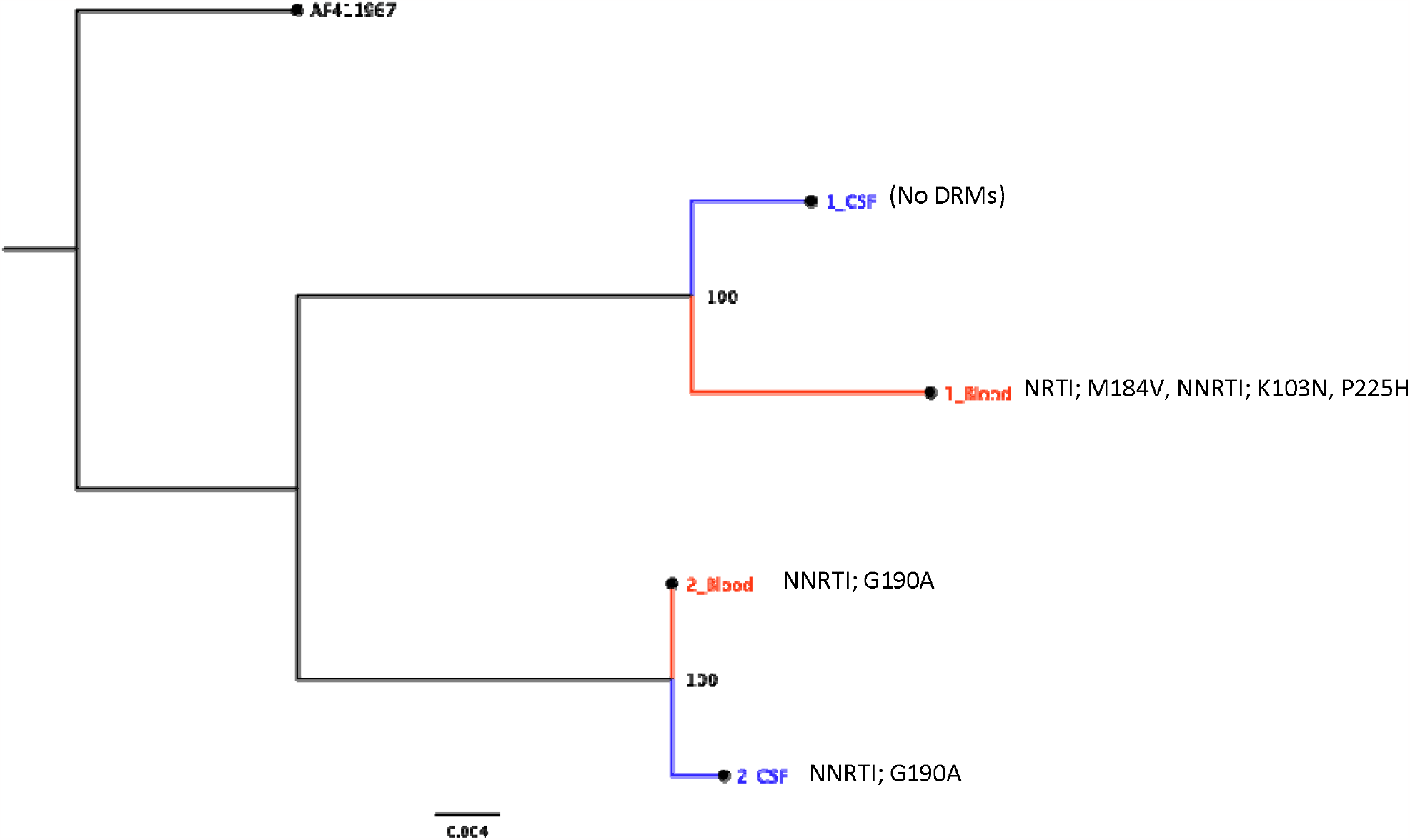
Maximum likelihood tree of the Reverse Transcriptase coding sequence (897 nucleotides) from 2 patients with detectable virus in blood and CSF. DRMs; drug resistance mutations, NRTI; nucleotide(side) reverse transcriptase inhibitor, NNRTI; non-nucleotide reverse transcriptase inhibitor.

### Brain imaging

Seven participants underwent brain MRI without contrast. Six brain MRIs showed FLAIR white matter hyperintensities in the cerebral hemispheres and one in addition had calcified foci in cerebral hemispheres, thought to be in keeping with past neurocysticercosis. This included 2 participants with detectable CSF VL and 4 without detectable CSF or plasma VL. One participant had an entirely normal brain MRI; this participant had an undetectable plasma VL but did not have a lumbar puncture.

### Adherence

Adherence was measured by random drug levels in plasma by mass spectrometry. Thirteen participants had drug levels measured in plasma, and 9 of them had drug levels measured in CSF. Drug levels were 2-3 log_10_ lower in CSF compared to plasma (Figure 3). Participants 1 and 2 (table 2) with detectable viruses in blood and CSF had drug levels below the limit of detection of the drug assays in both compartments (Figure 3).

**Figure 3:**
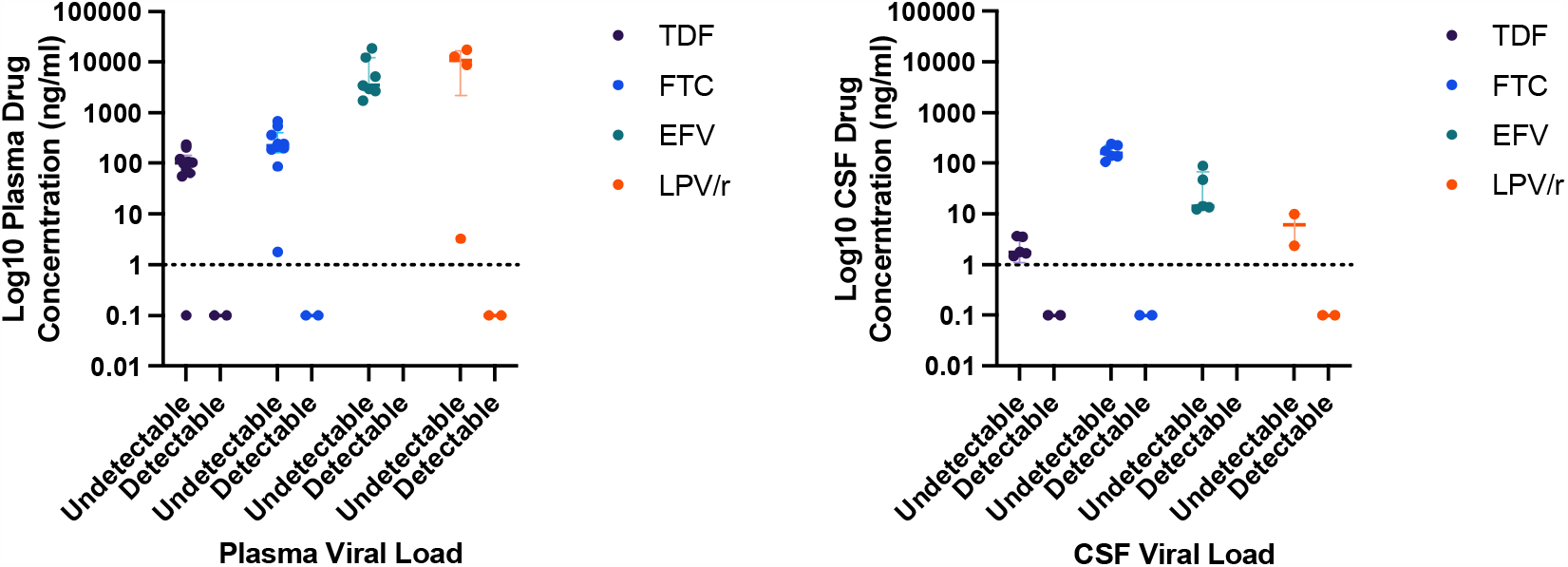
Antiretroviral drug levels. Nine participants with paired CSF and blood tenofovir (TDF), emtricitabine (FTC), efavirenz (EFV) and ritonavir boosted lopinavir (LPV/r) drug levels. Dotted line is limit of detection of assay. Undetectable levels were arbitrarily set at 0.1ng/ml.

## Discussion

We present the results at baseline for an ongoing longitudinal study, started pre-COVID and before the introduction of integrase inhibitor-based regimens. We discovered detectable HIV-1 VL in the CSF in 18.1% of participants with neurocognitive complaints on long term ART in an urban setting in South Africa. If applying the strict definition of CSF escape/discordance the prevalence is 9% and consistent with previously published prevalence from resource rich settings of up to 10%(10, 13, 14). A previous study in South Africa reported a CSF escape/discordance prevalence of 28%(20). However, the higher prevalence is likely due to the participants being recruited from an in-patient tertiary hospital setting, and therefore are likely to be sicker and not necessarily reflective of the broader population of PLWH in the community with mild-moderate neurocognitive complaints as represented by our study.

We discovered an association between detectable CSF HIV-1 VL and HIV-1 viraemia and furthermore this was associated with undetectable ART drug levels in both blood and CSF. In this cohort virological failure at baseline appears to be driven by poor ART adherence. This is a novel finding as previous studies exploring predictors of CSF escape have not measured drug levels in the CSF to determine the contribution of adherence. At baseline, the presence of detectable HIV-1 VL in the CSF was reflective of virological failure and probably due to poor ART adherence. This is an important finding because ART adherence is critical for durable viral suppression and also a modifiable factor that interventions could address. We also found an association with being on second line ritonavir boosted PI treatment. PI based combination therapy has previously been reported to be an independent predictor for CSF escape(13) and is thought to be due to poor CNS penetration of PIs(21), although adherence is likely also important.

Two participants had the virus population in their blood and CSF sequenced. These showed mutation in the NNRTI G190A in both CSF and blood in 1 participant and NNRTI K103N, P225H and NRTI M184V in the second participant in blood only. These mutations are reflective of exposure to previous NNRTI-based combination therapy. No minority variants were detected in NGS sequencing below 20%. Previous studies have shown CSF escape viruses with acquired resistant mutations (5, 8, 12, 13). However, it is as yet unknown if these emergent resistant viruses are the source of peripheral rebound on treatment interruption. We plan to follow up these participants to assess evolution of drug resistance in CSF escape HIV viruses and to explore whether these spill over into the blood.

Cognitive disorders continue to be reported in PLWH despite ART and undetectable HIV-1 in the blood. In this cohort, neurocognitive symptoms were identified on direct questioning. The IHDS screening test was less that 10 in 70% of participants, suggesting a potentially high prevalence of HAND in this setting. However, symptoms of depression occurred concurrently and may bias self-reported neurocognitive impairment. Interestingly headache was the most common symptom reported and it may very well be that headaches are an expression of stressful life events leading to depression. However, this needs to be explored further with a larger sample size.

Neuronal damage and inflammation as a result of HIV-1 replication in the CNS is implicated in HIV associated cognitive impairment and therefore clinical biomarkers for HIV-1 CSF replication are needed. We performed 3 Tesla brain MRI on a subset of participants. This did not discriminate between those with HIV-1 CSF replication and those without. This cohort had a low nadir CD4 at ART initiation, therefore the finding of punctate FLAIR white matter hyperintensities in the cerebral hemispheres in 6 out of 7 of participants including those without detectable HIV-1 in the CSF is likely to be a legacy of advanced HIV at ART initiation. Interestingly, one participant had a new diagnosis of calcified neurocysticercosis without a history of seizures. This is a prevalent condition in rural South Africa(22).

Our study contributes to the emerging field of HIV-1 CSF escape in SSA and its contribution to neurocognitive impairment. Our longitudinal study will determine if drug resistance mutations evolve in CNS and is a potential source of viral rebound. Recent initiation of dolutegravir based first-line combination ART in South Africa gives us the unique opportunity to study the evolution of integrase inhibitor resistance in the CNS. This of clinical importance in deciding optimal ART regimens and is a research priority for HIV cure efforts as HIV-1 CSF escape is a barrier to achieving lasting eradication of HIV from active and latent reservoirs.

## Data Availability

All data produced in the present study are available upon reasonable request to the authors

## Acknowledgements

We are thankful to the volunteers who participated in the study.

R.K.G is supported by Wellcome Senior Fellowship in Clinical Science (WT108082AIA).

D.A.C supported by Wellcome PhD Programme for Clinicians (206440/Z/17/Z).

The study was supported by The Rosetrees Trust.

## References

1. Sigal A, Baltimore D. As good as it gets? The problem of HIV persistence despite antiretroviral drugs. Cell Host Microbe. 2012;12(2):132–8.

2. Watters SA, Mlcochova P, Gupta RK. Macrophages: the neglected barrier to eradication. Curr Opin Infect Dis. 2013;26(6):561–6.

3. Lorenzo-Redondo R, Fryer HR, Bedford T, Kim EY, Archer J, Pond SLK, et al. Persistent HIV-1 replication maintains the tissue reservoir during therapy. Nature. 2016;530(7588):51–6.

4. Kugathasan R, Collier DA, Haddow LJ, El Bouzidi K, Edwards SG, Cartledge JD, et al. Diffuse White Matter Signal Abnormalities on Magnetic Resonance Imaging Are Associated With Human Immunodeficiency Virus Type 1 Viral Escape in the Central Nervous System Among Patients With Neurological Symptoms. Clin Infect Dis. 2017;64(8):1059–65.

5. Canestri A, Lescure FX, Jaureguiberry S, Moulignier A, Amiel C, Marcelin AG, et al. Discordance between cerebral spinal fluid and plasma HIV replication in patients with neurological symptoms who are receiving suppressive antiretroviral therapy. Clin Infect Dis. 2010;50(5):773–8.

6. Eden A, Marcotte TD, Heaton RK, Nilsson S, Zetterberg H, Fuchs D, et al. Increased Intrathecal Immune Activation in Virally Suppressed HIV-1 Infected Patients with Neurocognitive Impairment. PLoS One. 2016;11(6):e0157160.

7. Peluso MJ, Ferretti F, Peterson J, Lee E, Fuchs D, Boschini A, et al. Cerebrospinal fluid HIV escape associated with progressive neurologic dysfunction in patients on antiretroviral therapy with well controlled plasma viral load. AIDS. 2012;26(14):1765–74.

8. Collier DA, Haddow L, Brijkumar J, Moosa MS, Benjamin L, Gupta RK. HIV Cerebrospinal Fluid Escape and Neurocognitive Pathology in the Era of Combined Antiretroviral Therapy: What Lies Beneath the Tip of the Iceberg in Sub-Saharan Africa? Brain Sci. 2018;8(10).

9. Winston A, Antinori A, Cinque P, Fox HS, Gisslen M, Henrich TJ, et al. Defining cerebrospinal fluid HIV RNA escape: editorial review AIDS. AIDS. 2019;33 Suppl 2:S107–S11.

10. Eden A, Fuchs D, Hagberg L, Nilsson S, Spudich S, Svennerholm B, et al. HIV-1 viral escape in cerebrospinal fluid of subjects on suppressive antiretroviral treatment. J Infect Dis. 2010;202(12):1819–25.

11. Collier DA, Monit C, Gupta RK. The Impact of HIV-1 Drug Escape on the Global Treatment Landscape. Cell Host Microbe. 2019;26(1):48–60.

12. Mukerji SS, Misra V, Lorenz D, Cervantes-Arslanian AM, Lyons J, Chalkias S, et al. Temporal Patterns and Drug Resistance in CSF Viral Escape Among ART-Experienced HIV-1 Infected Adults. J Acquir Immune Defic Syndr. 2017;75(2):246–55.

13. Mukerji SS, Misra V, Lorenz DR, Uno H, Morgello S, Franklin D, et al. Impact of Antiretroviral Regimens on CSF Viral Escape in a Prospective Multicohort Study of ART-Experienced HIV-1 Infected Adults in the United States. Clin Infect Dis. 2018.

14. Di Carlofelice M, Everitt A, Muir D, Winston A. Cerebrospinal fluid HIV RNA in persons living with HIV. HIV Med. 2018;19(5):365–8.

15. Nightingale S, Geretti AM, Beloukas A, Fisher M, Winston A, Else L, et al. Discordant CSF/plasma HIV-1 RNA in patients with unexplained low-level viraemia. J Neurovirol. 2016;22(6):852–60.

16. Nightingale S, Dreyer AJ, Saylor D, Gisslén M, Winston A, Joska JA. Moving on From HAND: Why We Need New Criteria for Cognitive Impairment in Persons Living With Human Immunodeficiency Virus and a Proposed Way Forward. Clinical Infectious Diseases. 2021;73(6):1113–8.

17. Liu TF, Shafer RW. Web resources for HIV type 1 genotypic-resistance test interpretation. Clin Infect Dis. 2006;42(11):1608–18.

18. Katoh K, Standley DM. MAFFT multiple sequence alignment software version 7: improvements in performance and usability. Mol Biol Evol. 2013;30(4):772–80.

19. Minh BQ, Schmidt HA, Chernomor O, Schrempf D, Woodhams MD, von Haeseler A, et al. IQ-TREE 2: New Models and Efficient Methods for Phylogenetic Inference in the Genomic Era. Mol Biol Evol. 2020;37(5):1530–4.

20. Lustig G, Cele S, Karim F, Derache A, Ngoepe A, Khan K, et al. T cell derived HIV-1 is present in the CSF in the face of suppressive antiretroviral therapy. PLoS Pathog. 2021;17(9):e1009871.

21. Cusini A, Vernazza PL, Yerly S, Decosterd LA, Ledergerber B, Fux CA, et al. Higher CNS penetration-effectiveness of long-term combination antiretroviral therapy is associated with better HIV-1 viral suppression in cerebrospinal fluid. J Acquir Immune Defic Syndr. 2013;62(1):28–35.

22. Foyaca-Sibat H, Cowan LD, Carabin H, Targonska I, Anwary MA, Serrano-Ocana G, et al. Accuracy of serological testing for the diagnosis of prevalent neurocysticercosis in outpatients with epilepsy, Eastern Cape Province, South Africa. PLoS Negl Trop Dis. 2009;3(12):e562.

